# Inflammation-related Cognitive Risk Score (ICRS) as tool for predicting cognitive decline in older adults

**DOI:** 10.1101/2025.06.08.25329239

**Authors:** Suk Ling Ma, Ruonan Gao, Allen Ting Chun Lee, Linda Chiu Wa Lam

**Affiliations:** School of Arts and Humanities, Tung Wah College, Hong Kong; Department of Psychiatry, Faculty of Medicine, The Chinese University of Hong Kong, Hong Kong

**Keywords:** Inflammation, cognitive decline, risk prediction

## Abstract

**Objectives:** The prevalence of dementia increased exponentially with the aging population globally. Early detection of cognitive decline at prodromal stage allows room for intervention to prevent or delay the progress of dementia. This study aimed to identify a simple and accurate blood test to identify older adults who are at high risk of cognitive decline.

**Methods:** Inflammation-related marker panel was derived from gene expression data from Gene Expression Omnibus (GEO) database and Inflammation-related Cognitive Risk Score (ICRS) was constructed to predict the risk of cognitive decline. 142 non-demented older adults from the community were recruited and followed-up for two years. RT-qPCR was performed for genes included in the marker panel using blood samples collected at baseline and ICRS was calculated for each older adults. Cognitive assessments were performed at baseline and 2-year follow-up and ICRS was used to identify older adults who were at high risk of cognitive decline.

**Results:** ICRS was significantly higher in older adults without cognitive decline when compared to those with cognitive decline (p=0.011). When the ICRS was further adjusted by vascular factors as ICRS-adj, the differentiation was significantly improved (p<0.001) with increased sensitivity and specificity.

**Conclusion:** Our result showed that ICRS-adj can predict cognitive decline with high sensitivity and specificity and this will be beneficial to identify older adults who are at high risk of dementia for early intervention.

## Introduction

Early detection of cognitive decline is critical in public health. With the increase in aging population and average lifespan, the prevalence of dementia is expected to increase exponentially. Over 55 million people suffered from dementia worldwide and the number is expected to be doubled in 20 years. In Hong Kong, a recent epidemiology study reported the prevalence of mild neurocognitive disorders (including mild cognitive impairment (MCI)) is up to 21.8% (1). Studies showed that intervention is effective at prodromal stage of dementia to slow down or reduce the progression to dementia. Some biomarkers such as positron emission tomography (PET), amyloid imaging and cerebrospinal fluid (CSF) tau/amyloid beta (Aβ) may be useful for early detection but they are either invasive or expensive. On the other hand, a simple and inexpensive blood test that can identify the high risk individual of cognitive decline will serve as an excellent tool in community for early detection and intervention for dementia.

The association between inflammation and Alzheimer’s disease (AD) was supported by different lines of evidence. Inflammatory response was observed in postmortem AD patients (2). A recent meta-analysis study included more than 70 studies showed inflammatory cytokines such as as interleukin-1beta (IL-1β), IL-6 and and tumor necrosis factor-α (TNF-α) were increased in both AD and MCI. They also showed IL-6 was associated to cognitive decline (3). Another study further showed low-grade systemic inflammation was associated to various markers related to AD (4). More importantly, studies showed the inflammatory profile changed before the onset of symptoms (5), suggesting inflammatory markers might be useful for early detection of cognitive decline.

In this study, we aimed to identify inflammation-related marker panel and construct a risk score, Inflammation-related Cognitive Risk Score (ICRS) to identify older adults who are at high risk of cognitive decline.

## Method

### Participants

142 non-demented older adults from the community were recruited and they were screened by our trained research staff for eligibility. The inclusion criteria were aged 60-85, ethnic Chinese, living in the community, no dementia (Clinical Dementia Rating (CDR) 0 or 0.5) and free of depressive symptoms and no neurological conditions that may affect cognition. During recruitment, our research staff explained the procedure and obtained informed consent from the participants. The study was approved by the Clinical Research Ethics Committee of the Chinese University of Hong Kong.

### Assessments

All participants underwent cognitive assessments with CDR and Hong Kong Montreal Cognitive Assessment (HK-MoCA) (6,7). HK-MoCA is a locally validated cognitive screening test sensitive in detecting early memory and executive deficits. The total score of HK-MoCA is 30, the higher the score, the better the cognition. The older adults received baseline and 2-year follow-up assessment to compare the cognitive performance over 2 years.

Venous blood samples at baseline assessment were collected in anticoagulant-free tubes in the morning to minimize diurnal variation of gene expression and allow measurements of overnight cholesterol (high-density lipoprotein (HDL) and low-density lipoprotein (LDL)) and blood glucose levels. In addition, ApoE genotyping was performed as described elsewhere.

### Inflammation-related marker panel and Gene expression quantification

Gene expression data was obtained from Gene Expression Omnibus (GEO) database (GSE63063) and the dataset contained normal controls and Alzheimer’s disease (AD). Differentially expressed genes (DEGs) were identified by a linear regression model using the R/Bioconductor *limma* package. Genes with log fold change > 1 and Benjamini–Hochberg– adjusted p < 0.05 were considered as significant DEGs. 40 DEGs involved in inflammatory pathways were selected to form the inflammation-related marker panel.

PMBC was isolated from the collected blood samples after centrifugation and was stabilized by Trizol. The samples were stored at -80C until RNA extraction. RNA was extracted from Trizol stabilized blood samples and converted to cDNA by reverse transcription. Real-time quantitative PCR (RT-qPCR) combined with TaqMan probes was used to quantify the expression level of genes in the inflammatory marker panel using LightCycler 480 (Roche). Fold changes were calculated by comparing the Ct values of the target gene and the internal control gene (house-keeping gene) using the ΔΔCt method.

### Risk score construction and Statistical analysis

Our team’s earlier study suggested 1-standard deviation (SD) drop in cognitive assessment, Chinese version of Mini-Mental State Examination (MMSE) was highly sensitive in predicting the development of dementia in 5 years (8). Therefore 1-SD drop in HK-MoCA was regarded as predictor for future cognitive decline (outcome measurement). The gene expression was compared between older adults with 1-SD drop in HK-MoCA and those without drop by students’ t-test to identify significant genes. These genes were selected to construct the Inflammation-related Cognitive Risk Score (ICRS). The score was constructed based on Cox regression model and was calculated according to the equation: risk score = ∑ coefficient value *gene expression level. Based on this calculation, ICRS was calculated for each subject and the ICRS was compared between older adults with cognitive decline and those without by students’ t-test. All statistical analysis were done by either R or SPSS 29.0.

## Results

### Characteristics of the cohort

142 non-demented older adults were recruited for this study and the mean age was 69.5 (SD =5.6) and 59.2% of them were females. At baseline, 29.6% and 70.4% of the participants were rated as CDR 0 and 0.5 respectively (Table 1). No significance difference in age and gender for participants with different CDR at baseline. The HK-MoCA score for older adults of CDR 0 was slightly higher than those of CDR 0.5, which is expected but the difference did not reach statistical significance. At 2-year follow-up, 16.9% of the participants showed cognitive decline (as defined by 1 SD drop in HK-MoCA) with over 60% of them were CDR 0.5 at baseline.

**Table 1.**
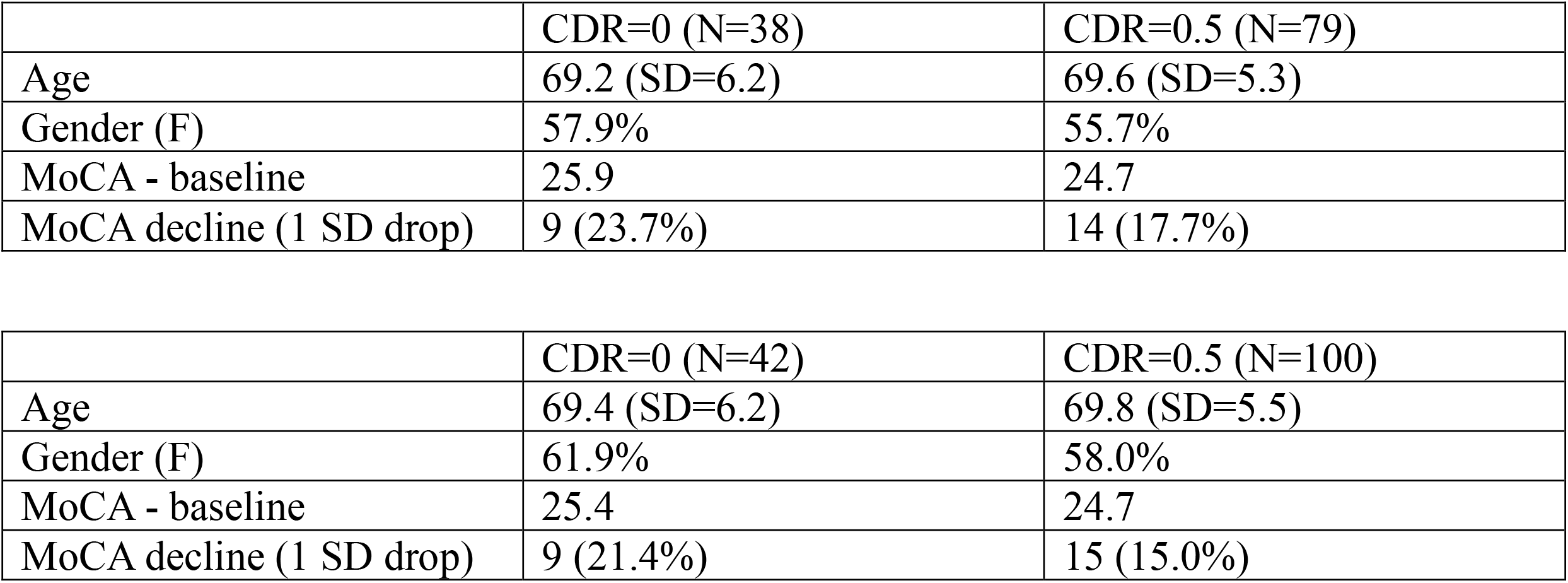
Table showing the demographic characteristics of the older adults of the study.

### Inflammation-related Cognitive Risk Score (ICRS) construction and association to cognitive decline

Gene expression data from GSE63063 was downloaded and analysed by R package. 2609 DEGs were identified. Genes related to inflammation were further selected and analysed to form the inflammation marker panel. Gene expression of the components of the inflammation-related marker panel was quantified by RT-qPCR. ICRS was constructed based on Cox regression model and the score was calculated for each subject accordingly. The ICRS was significantly higher in older adults without cognitive decline when compared to those with cognitive decline (Figure 1) (p=0.011). Previous studies suggested cardiovascular risk factors played an important role in the risk of dementia, ratio of HDL/total cholesterol and LDL cholesterol were taken into account for the calculation of ICRS. When these two parameters exceeded the recommended range (i.e. above 5 for HDL/total cholesterol and above 4.5 mmol/L for LDL cholesterol), the median of the ICRS was added to adjust and this new score was termed as ICRS-adjusted (ICRS-adj). Using ICRS-adj, the differentiation between older adults who might have cognitive decline over 2 years and those who might not was greatly improved, with a clear cutoff (Figure 2) (p<0.001).The Receiver Operating Characteristic (ROC) curve compared the performance of ICRS and ICRS-adj and it showed the ICRS-adj outperformed the original score and yielded 0.922 (Figure 3).

**Figure 1.**
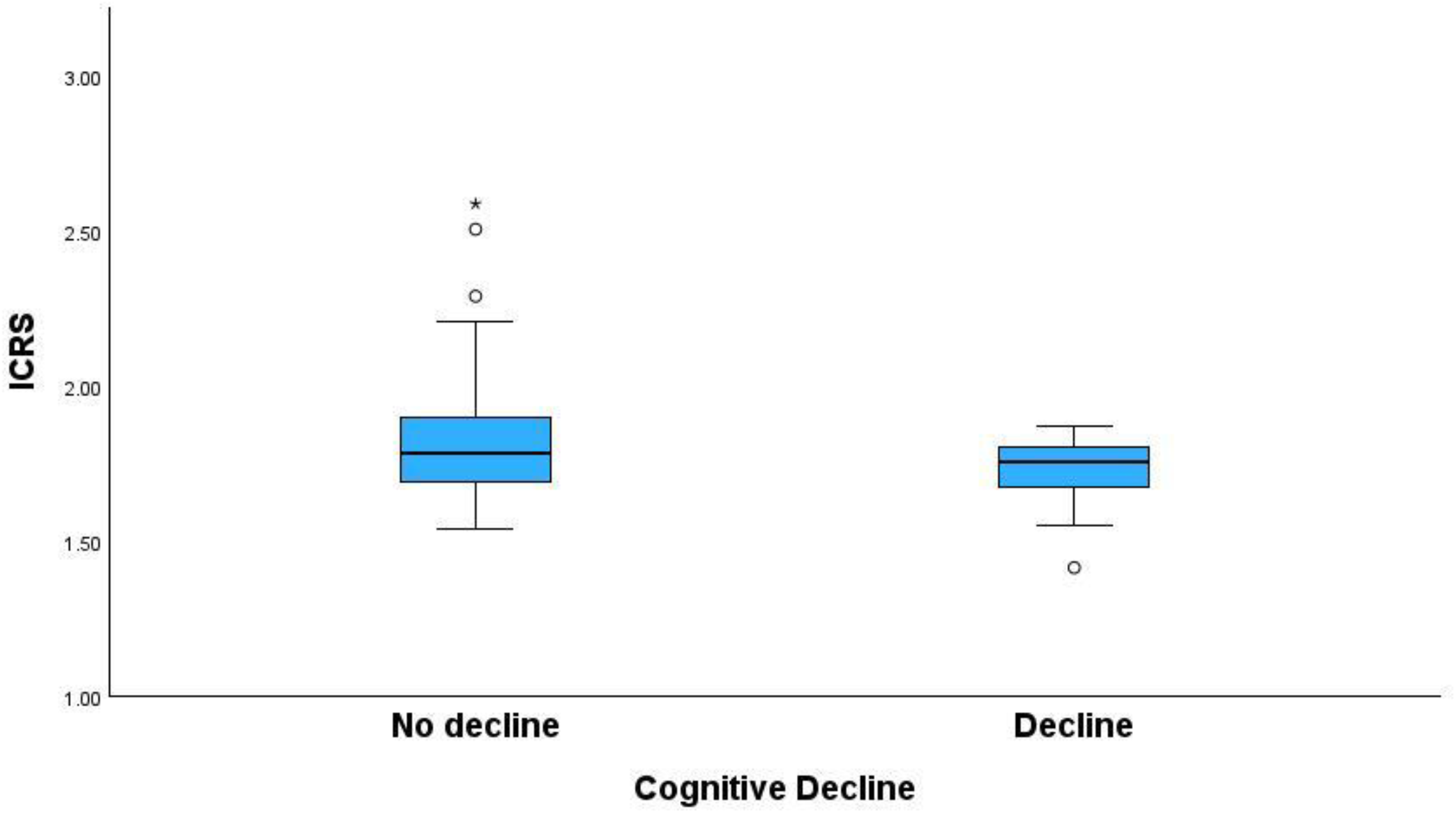
Boxplot showing ICRS can differentiate between older adults without cognitive decline and those with cognitive decline.

**Figure 2.**
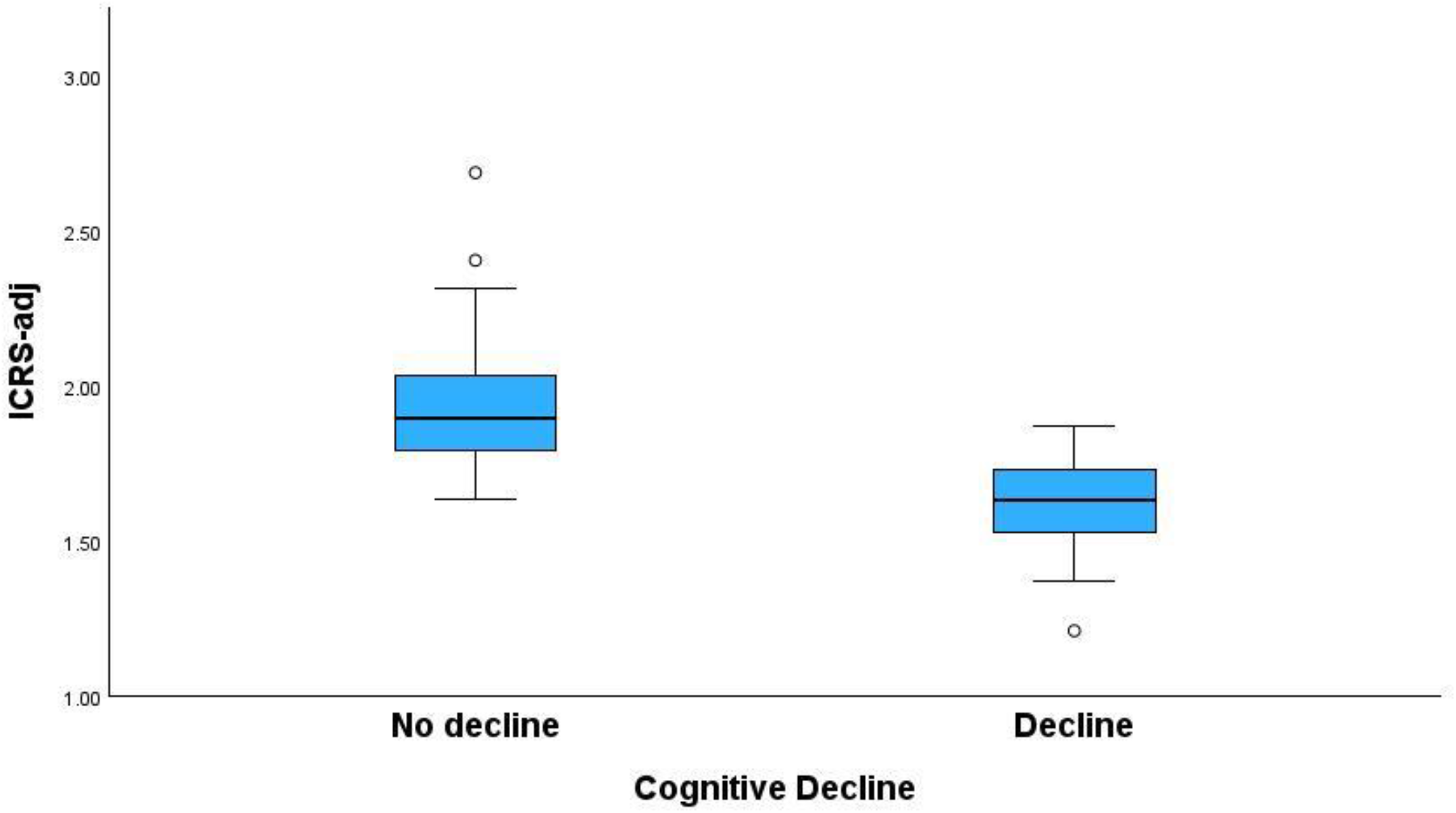
Boxplot showing ICRS-adj can differentiate between older adults without cognitive decline and those with cognitive decline.

**Figure 3.**
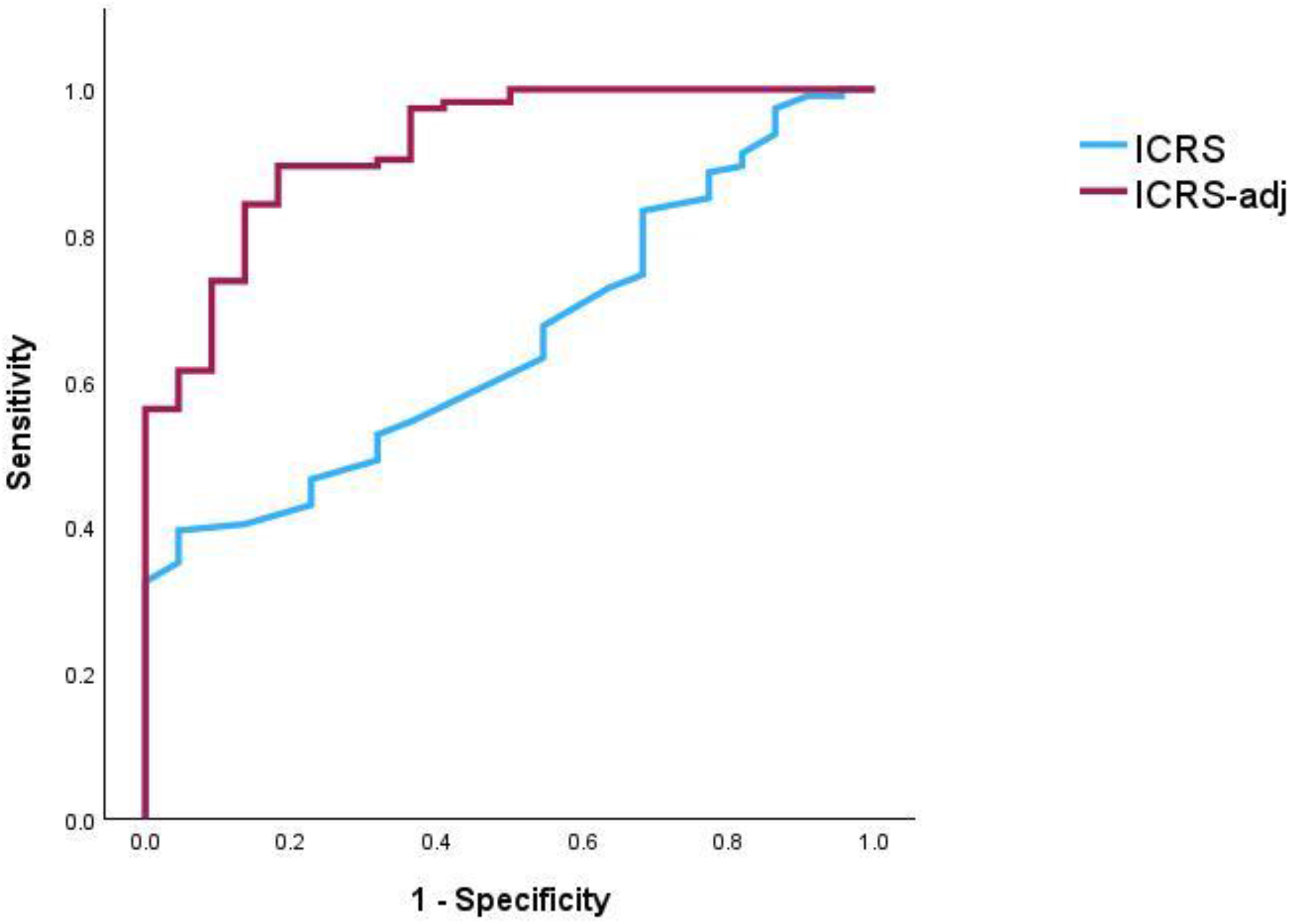
Receiver operating characteristic (ROC) curves showing the performance of ICRS and ICRS-adj.

## Discussion

Over the past decade, there were many studies trying to identify biomarkers for the diagnosis of AD (9–11) or predict the risk of AD (12–16). In 2024, the Alzheimer’s Association Workgroup from The National Institute on Aging and the Alzheimer’s Association revised the criteria for diagnosis and staging of AD but it still regarded amyloid β (Aβ) and p-tau as the gold standard, with the inclusion of glial fibrillary acidic protein (GFAP), a protein involved in astrocytic activation (inflammation) (17). Nowadays, there are monoclonal antibodies (aducanumab, lecanemab and donanemab) for AD which served as promising therapies (18). However, the possible side-effects such as amyloid-related imaging abnormalities (ARIA) and the high cost of medication made it not an option for everyone. On the other hand, studies suggested that many risk factors for dementia are modifiable (19), therefore there is pressing need for a simple and effective tool to identify individuals who are at high risk of cognitive decline.

In this study, cognitive decline was defined by 1-SD drop (equivalent to 2 points in the current study) in HK-MoCA. MoCA is a widely used cognitive assessment to screen for MCI and it is validated in different languages including HK-MoCA (6,7). Our group and others used 1-SD or 2 points drop in MoCA or MMSE as cutoff for cognitive decline (8,20), that our previous finding suggested that this was highly sensitive in predicting future chance of dementia. In addition, we identified an inflammatory-related marker panel from GEO database, focusing on genes which may implicate in the inflammatory. The rationale of choosing inflammatory-related genes is the close relationship between inflammation and the pathogenesis of AD (2,4). ICRS was further developed based on the regression model and our results showed that ICRS was significantly lower in older adults with cognitive decline. Cardiovascular risk factors are regarded as important risk factors for dementia (21,22), therefore these parameters were further incorporated with ICRS as ICRS-adj. Our finding showed that the sensitivity and specificity of ICRS-adj improved significantly.

Previous studies investigated the association of inflammatory markers and the risk of cognitive decline or dementia and the results were controversial. The diagnosis of dementia was regarded as the outcome in several studies and this might weaken the predictive or beneficial value of the inflammatory markers (23,24). On the other hand, some studies were able to predict cognitive decline using inflammatory markers (9,13,25). The Lancet Commission on dementia suggested that there are 14 potentially modifiable risk factors for dementia and half of the risk of dementia might be delayed or reduced (19). ICRS-adj incorporated one of the potentially modifiable risk factors and it is calculated base on the inflammatory marker panel which might reflect inflammation caused by other modifiable risk factors as mentioned. Our study result showed that ICRS-adj can predict cognitive decline with high sensitivity and specificity and this will be beneficial to identify older adults who are at high risk of dementia for early intervention. There are some limitations for the study including small sample size and short follow-up duration. Future studies will be required to validate the findings. In summary, our findings suggested that ICRS-adj might be feasible for early screening of cognitive decline and provide a window for early intervention.

## Data Availability

All data produced in the present study are available upon reasonable request to the authors

## Acknowledgement

This study was funded by the Health and Medical Research Fund (HMRF) (Ref no. 09200246) and HMRF Commissioned Research (Ref no. MHS-P1(Part3)-CUHK), Health Bureau, Hong Kong SAR Government.

## Reference

1. Lam LCW, Chan WC, Lee ATC et al. The Hong Kong Mental Morbidity Survey for Older People – HKMMSOP. Hong Kong.

2. Gomez-Nicola D, Boche D. Post-mortem analysis of neuroinflammatory changes in human Alzheimer’s disease. Alzheimers Res Ther 2015; 7: 42.

3. Leonardo S, Fregni F. Association of inflammation and cognition in the elderly: A systematic review and meta-analysis. Front Aging Neurosci 2023; 15: 1069439.

4. Valletta M, Vetrano DL, Rizzuto D et al. Blood biomarkers of Alzheimer’s disease in the community: Variation by chronic diseases and inflammatory status. Alzheimers Dement 2024; 20: 4115–4125.

5. Heppner FL, Ransohoff RM, Becher B. Immune attack: the role of inflammation in Alzheimer disease. Nat Rev Neurosci 2015; 16: 358–372.

6. Yeung PY, Wong LL, Chan CC, Leung JL, Yung CY. A validation study of the Hong Kong version of Montreal Cognitive Assessment (HK-MoCA) in Chinese older adults in Hong Kong. Hong Kong medical journal = Xianggang yi xue za zhi 2014; 20: 504–10.

7. Nasreddine ZS, Phillips NA, Bédirian V et al. The Montreal Cognitive Assessment, MoCA: a brief screening tool for mild cognitive impairment. J Am Geriatr Soc 2005; 53: 695–699.

8. Wong CHY, Leung GTY, Fung AWT, Chan WC, Lam LCW. Cognitive predictors for five-year conversion to dementia in community-dwelling Chinese older adults. Int Psychogeriatr 2013; 25: 1125–1134.

9. Jiang Y, Uhm H, Ip FC et al. A blood-based multi-pathway biomarker assay for early detection and staging of Alzheimer’s disease across ethnic groups..

10. Park YH, Hodges A, Simmons A et al. Association of blood-based transcriptional risk scores with biomarkers for Alzheimer disease. Neurol Genet 2020; 6: e517.

11. Barthélemy NR, Salvadó G, Schindler SE et al. Highly accurate blood test for Alzheimer’s disease is similar or superior to clinical cerebrospinal fluid tests. Nat Med 2024; 30: 1085–1095.

12. Chaudhury S, Brookes KJ, Patel T et al. Alzheimer’s disease polygenic risk score as a predictor of conversion from mild-cognitive impairment. Translational Psychiatry 2019; 9: 1– 7.

13. Kipinoinen T, Toppala S, Rinne JO, Viitanen MH, Jula AM, Ekblad LL. Association of Midlife Inflammatory Markers With Cognitive Performance at 10-Year Follow-up. Neurology 2022; 99: e2294–e2302.

14. Guduguntla BA, Vasbinder A, Anderson E et al. Biomarkers of chronic inflammation and cognitive decline: A prospective observational study. Alzheimer’s & Dementia: Diagnosis, Assessment & Disease Monitoring 2024; 16: e12568.

15. Wang X, Bakulski KM, Karvonen-Gutierrez CA et al. Blood-based biomarkers for Alzheimer’s disease and cognitive function from mid-to late life. Alzheimers Dement 2023; 20: 1807–1814.

16. Chi GC, Fitzpatrick AL, Sharma M, Jenny NS, Lopez OL, DeKosky ST. Inflammatory Biomarkers Predict Domain-Specific Cognitive Decline in Older Adults. The Journals of Gerontology: Series A 2017; 72: 796–803.

17. Jack Jr. CR, Andrews JS, Beach TG et al. Revised criteria for diagnosis and staging of Alzheimer’s disease: Alzheimer’s Association Workgroup. Alzheimer’s & Dementia 2024; 20: 5143–5169.

18. Kim B-H, Kim S, Nam Y, Park YH, Shin SM, Moon M. Second-generation anti-amyloid monoclonal antibodies for Alzheimer’s disease: current landscape and future perspectives. Transl Neurodegener 2025; 14: 6.

19. Livingston G, Huntley J, Liu KY et al. Dementia prevention, intervention, and care: 2024 report of the Lancet standing Commission. The Lancet 2024; 404: 572–628.

20. Zuo L, Dong Y, Liao X et al. Risk factors for decline in Montreal Cognitive Assessment (MoCA) scores in patients with acute transient ischemic attack and minor stroke. J Clin Hypertens (Greenwich) 2022; 24: 851–857.

21. Kjeldsen EW, Frikke-Schmidt R. Causal cardiovascular risk factors for dementia: insights from observational and genetic studies. Cardiovasc Res 2025; 121: 537–549.

22. Jia R, Wang Q, Huang H, Yang Y, Chung YF, Liang T. Cardiovascular disease risk models and dementia or cognitive decline: a systematic review. Front Aging Neurosci 2023; 15: 1257367.

23. Fang Y, Doyle MF, Chen J et al. Association between inflammatory biomarkers and cognitive aging. PLoS ONE 2022; 17: e0274350.

24. Bocharova M, Borza T, Watne LO et al. The role of plasma inflammatory markers in late-life depression and conversion to dementia: a 3-year follow-up study. Mol Psychiatry 2025;

25. Thomas A, Guo J, Reyes-Dumeyer D et al. Inflammatory biomarkers profiles and cognition among older adults. Sci Rep 2025; 15: 2265.

